# Biologically informed instrument selection for dietary Mendelian randomization using chemosensory receptor variants

**DOI:** 10.64898/2026.02.05.26345702

**Authors:** Liang-Dar Hwang, Cailu Lin, David M Evans, Nicholas G Martin, Danielle R Reed, Paule V Joseph

**Author notes:** Correspondence: Liang-Dar Hwang, Danielle R. Reed, or Paule V. Joseph.

## Abstract

**Background:** Mendelian randomization (MR) is increasingly used for causal inference in nutritional epidemiology; however, dietary MR studies often rely on instruments statistically selected from genome-wide association studies of self-reported intake, which are vulnerable to pleiotropy and reverse causation and may violate core MR assumptions. We aimed to develop and evaluate a biologically informed framework for selecting valid genetic instruments for dietary exposures, based on genes encoding taste and olfactory receptors that mediate chemosensory inputs and shape food preferences and dietary behaviour.

**Methods:** We prioritised 1,214 nonsynonymous variants in 30 taste and 295 olfactory receptor genes with minor allele frequency ≥1%. Associations with 140 food-liking traits were tested in UK Biobank participants aged 37 to 73 years. Candidate variants were evaluated using a multi-stage filtering pipeline designed to improve instrument validity. This included replication in an independent younger cohort (Avon Longitudinal Study of Parents and Children, age 25), concordance between food liking and intake, exclusion of associations with socioeconomic status, assessment of food specificity accounting for linkage disequilibrium and co-consumption patterns, and directionality testing to reduce reverse causation. Retained variants were applied as instruments in MR analyses to assess cardiometabolic outcomes.

**Results:** We identified 268 nonsynonymous variants within 101 olfactory and 16 taste receptor genes associated with 96 food-liking traits. The filtering process yielded 28 candidate instruments for 24 foods. Among these, the instrument for onion liking uniquely satisfied all criteria for classification as high confidence. To demonstrate clinical relevance, genetically proxied onion liking was associated with lower blood pressure and a reduced risk of type 2 diabetes in MR analyses, with no evidence of effects on body mass index, glycaemic traits, or serum lipid levels.

**Conclusions:** Guiding genetic instrument selection using chemosensory receptor genes provides a biologically informed strategy for dietary Mendelian randomization that reduces susceptibility to pleiotropy and reverse causation. This framework enables more robust causal evaluation of diet–disease relationships and strengthens inference in nutritional epidemiology and public health research.

## Introduction

Unhealthy diets are a major contributor to the global disease burden, accounting for an estimated 11 million premature deaths annually, primarily from cardiovascular disease, followed by cancer, and type 2 diabetes ^1^. Rates of early-onset obesity, cancers ^2^, and diabetes ^3^ continue to rise, placing increasing pressure on healthcare systems. Despite extensive observational evidence linking diet to chronic disease, robust causal evidence for specific foods, dietary patterns, or nutrients remains limited. Randomized controlled trials (RCTs), the gold standard for causal inference, are rarely feasible in nutrition due to long follow-up periods, high costs, logistical constraints, and poor adherence ^4^. Observational studies are frequently confounded or affected by reverse causation; for example, while observational studies suggested that vitamin E supplementation lowers coronary artery disease risk, large-scale RCTs failed to replicate this effect ^5^. Such discrepancies underscore the need for rigorous methods that can reliably identify the causal effects of diet.

Mendelian randomization (MR) provides an alternative framework for causal inference. By using genetic variants as instrumental variables (IVs), MR mimics the randomization process of RCTs through Mendel’s Laws of Segregation and Independent Assortment, thereby reducing confounding ^6^. MR has reshaped the understanding of cardiometabolic disease by confirming the causal role of low-density lipoprotein cholesterol in cardiovascular disease, while demonstrating that associations with high-density lipoprotein cholesterol are likely non-causal. These findings align with RCTs and have influenced clinical practice and drug development ^7^. Because MR reflects lifelong exposure effects, it offers practical advantages over short-term, resource-intensive RCTs.

The application of MR in nutrition research is expanding ^8^, with studies clarifying the causal effects of coffee ^9^, alcohol ^10^, and milk intake ^11^. Despite these advances, MR analyses of foods and dietary patterns remain challenging ^8^, mainly because it is difficult to identify valid IVs. The MR exclusion restriction assumption requires that genetic variants affect outcomes solely through the exposure (i.e., no horizontal pleiotropy; **Figure 1**, blue arrow). Many dietary instruments are selected solely based on statistical significance in genome-wide association studies (GWAS), thereby increasing susceptibility to pleiotropy. This problem is compounded by the fact that most diet GWAS have been conducted in older adults ^12–21^, whose food choices may reflect pre-existing health conditions (**Figure 1**, orange arrow). For example, *FTO* variants associated with higher body mass index ^22^ are counterintuitively related to decreased sugar intake ^13^, and *APOE* variants linked to increased risk of cardiovascular ^23^ and neurological disorders ^24^ are likewise associated with reduced preference for fried foods ^19^. These patterns are unlikely to reflect primary effects on diet; instead, they suggest reverse causation, in which individuals modify their diet after diagnosis ^14,15^. Using such loci as IVs in MR would introduce substantial bias and exacerbate inconsistency in nutritional epidemiology.

**Figure 1:**
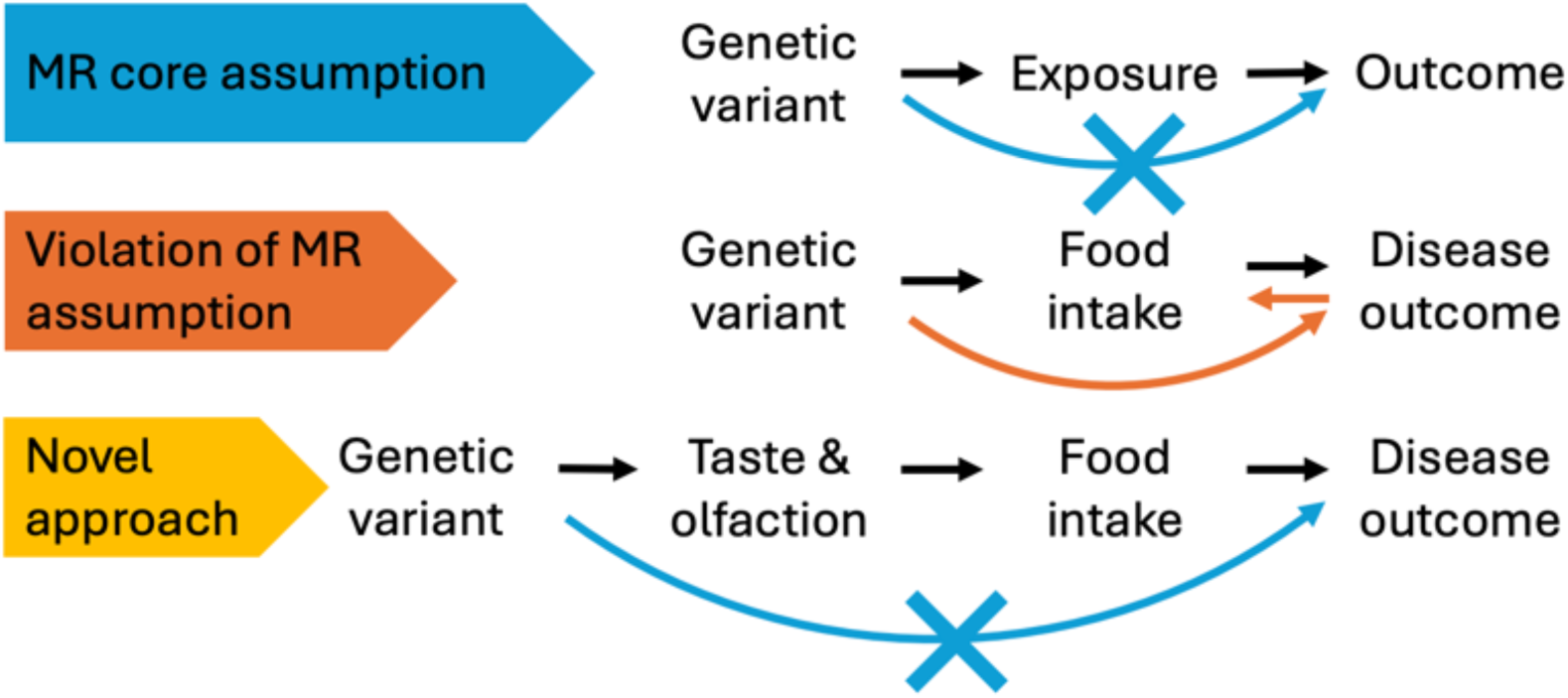
Genetic variants associated with chemosensory perception as novel instrumental variable for dietary exposures in Mendelian randomization (MR) for causal inference. MR assumes that a genetic variant affects an outcome solely through the exposure of interest (i.e., no pleiotropy; **Top**, blue arrow). However, most variants associated with dietary intake have been identified in older individuals, where food choices often reflect pre-existing health conditions (**Middle**, orange arrow) rather than cause them. The effect of genetic variants associated with chemosensory perception on diet is less likely to be influenced by health status, making them superior instruments for dietary exposures (**Bottom**).

Genetic variants that influence taste and olfactory perception show promise as IVs for dietary exposures. The chemical senses play a central role in shaping food preferences and choices, and variants associated with chemosensory perception ^25–30^ have consistently been linked to differences in dietary behavior ^31–34^. For example, variants in bitter taste receptor genes influence the bitterness of quinine ^26^ and caffeine ^28^ and predict the consumption of bitter beverages ^31^. Recent large-scale GWAS of food liking and intake have also identified variants within taste and olfactory receptor genes ^14,18,19^. Because chemosensory perception is a key determinant of eating behaviour—well before the onset of most chronic disease ^35^—associations between these variants and dietary intake are less susceptible to socioeconomic or health-related confounding. They are less likely to reflect disease-driven behavioral changes. These characteristics make genetic variants in chemosensory pathways biologically credible candidates for use as IVs in dietary MR.

Here, we evaluated whether genetic variants in taste and olfactory receptor genes can serve as IVs for dietary exposures. We prioritized nonsynonymous variants within chemosensory receptor genes (i.e., the taste receptor type 1 [T1Rs] and type 2 [T2Rs] families, and the olfactory receptor [OR] gene families), as these variants alter amino acid sequences and are therefore more likely to affect receptor structure and potentially ligand binding. Using UK Biobank food-liking traits ^36^, we identified candidate variants. We applied stringent filtering criteria: replication in the younger Avon Longitudinal Study of Parents and Children (ALSPAC) cohort ^37,38^, concordant associations with intake of the same foods, lack of association with socioeconomic status, and evidence of specificity to a single food after accounting for linkage disequilibrium (LD). We then used these instruments to perform MR analyses of the effects of food liking on cardiometabolic traits. An overview of the study design is presented in **Supplementary Figure 1**.

## Methods

### Sample and genotyping

UK Biobank is a prospective cohort study of ∼500,000 participants (aged 37–73 years; 54.4% female; ∼5% of those invited) recruited across England, Wales, and Scotland between 2006 and 2010. Baseline questionnaires, clinical assessments and biospecimens have been described previously ^36^. This project used the resource under Application 57780. Genotyping was performed using the Affymetrix UK BiLEVE Axiom or Affymetrix UK Biobank Axiom arrays. Imputation was performed using IMPUTE2 software with the UK10K haplotype and Haplotype Reference Consortium reference panels ^36^. Single nucleotide polymorphisms (SNPs) with a call rate < 90%, minor allele frequency < 0.005, imputation score < 0.3, and Hardy–Weinberg equilibrium score of *P* < 1.0 × 10^-6^ were excluded. Ancestry was assigned using a distance-based genetic relationship and fingerprinting (Graf) method ^39^; individuals clustering with the European group were retained. Relatedness was defined as having a KING kinship coefficient > 0.0884 (i.e., third-degree relatives or closer) ^40^; one individual from each pair was removed.

ALSPAC is a longitudinal birth cohort study that enrolled pregnant women residing in and around the city of Bristol in the South West of England with expected delivery dates between April 1, 1991, and December 31, 1992 ^37,38^. The enrolled cohort included 15,247 pregnancies resulting in 14,775 live-born babies. The mothers and their children have been followed up through postal questionnaires and at clinics. A searchable variable dictionary is available online (http://www.bristol.ac.uk/alspac/researchers/our-data/). The ALSPAC Ethics and Law Committee and local Research Ethics Committees granted ethical approval. This study used the children’s cohort. Genotyping was performed using the Illumina HumanHap550 quad chip array. SNPs with a call rate < 90%, minor allele frequency < 0.01, and Hardy–Weinberg equilibrium score of *P* < 1.0 × 10^-6^ were excluded. Imputation was performed using Impute V2.2.2 with the 1000 Genomes Phase 1 Version 3 reference panel ^38^. Ancestry was assessed using multidimensional analysis and compared with HapMap II (release 22) European descent (CEU), Han Chinese, Japanese, and Yoruba reference populations; all individuals with non-European ancestry were removed. Relatedness was defined using a genome-wide identity-by-descent estimated from PLINK ^41^; one individual from each pair with PI_HAT > 0.10 was removed.

### Common missense, nonsense, and frameshift variants in chemosensory receptor genes

We downloaded the complete list of human genes (GRCh37/hg19) from the GENECODE project^42^ and extracted taste receptor genes (i.e., gene symbol starting with *TAS1R* or *TAS2R*) and olfactory receptor genes (i.e., gene symbol starting with *OR*). We excluded pseudogenes and genes without chromosome or base-pair information. Based on the chromosome locations of each gene, we extracted all variants within each gene and excluded those with a minor allele frequency < 0.01 in the UK Biobank. The remaining genetic variants were annotated using Haploreg (v4.2) ^43^ and dbSNP ^44^ for functional annotations. Only variants annotated as missense, nonsense, or frameshift mutations were included in the analysis.

### Food liking and food intake

Food-liking traits from UK Biobank were collected through an online questionnaire comprising 152 items, including 140 food and drink items and additional non-food items that captured liking for health-related behaviors such as physical activity (https://biobank.ndph.ox.ac.uk/showcase/showcase/docs/foodpref.pdf). Participants rated their liking on a 9-point Hedonic scale, with 1 corresponding to “Extremely dislike” and 9 to “Extremely like”. The questionnaire was administered in 2019 to all UK Biobank participants who had agreed to be recontacted by the study. Participants who answered “Never tried” or “Do not wish to answer“ were excluded. Data from up to 182,165 individuals were available for analysis. For significant SNP-food-liking associations, we further examined the associations between the same SNPs and the intake of the same food items. Dietary intake in the UK Biobank was assessed using i) a touchscreen dietary frequency questionnaire from all participants during their visits to the assessment center and ii) a follow-up online 24-hour recall dietary questionnaire ^45^ from up to 214,950 individuals. Answers of “None” from the 24-hour recall were recoded to 0. Data collected from multiple instances were averaged before analysis.

Data on food liking in ALSPAC were collected from children of pregnant women recruited to the study when they were 25 years old using the Life at 25+ Questionnaire. Preferences for 97 food items were measured using a 9-point Hedonic scale similar to the one used in UK Biobank, with 1 corresponding to “Extremely dislike”, 9 to “Extremely like”, and 0 to “Never tasted”. From October 2017, participants received an email containing a username/password for the online version of the questionnaire. Participants without email addresses or who did not respond to the email were sent a paper questionnaire. The questionnaire was sent out to 10,001 live-born children within the ALSPAC cohort. As of 1^st^ October 2018, 4398 completed questionnaires had been returned. Only unrelated individuals of European ancestry with genotyping data, age, and sex information, and who provided an answer between 1 and 9 were included, leaving a total of up to 2,779 responses per food item, with 64% female. The ALSPAC participants included in this study did not overlap with those from the UK Biobank.

### Statistical analysis

We first calculated the effect of each SNP on each food-liking trait in the UK Biobank using a linear regression model. Covariates included sex, age, and the first 10 genetic principal components (PCs) (Formula 1).

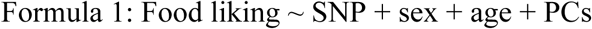

A prior significance threshold was set at an FDR-adjusted p-value of 0.05 to account for multiple testing. Four downstream analyses were performed following the significant associations.

First, for significantly associated SNPs, we assessed their associations with socioeconomic status, proxied by the Townsend Deprivation Index, in the UK Biobank using GWAS summary statistics from the OpenGWAS database ^46^.

Second, for SNPs identified in the UK Biobank, we investigated their association with their corresponding food-liking traits in the younger ALSPAC cohort using the same linear regression model. For SNPs unavailable in ALSPAC, their proxies with a LD of r^2^ (square of the Pearson correlation coefficient between allele dosages of two SNPs) > 0.8 in the British in England and Scotland (GBR) population were identified using the LDproxy Tool ^47^. We performed a heterogeneity test to compare the difference in SNP effects between the UK Biobank and ALSPAC.

Third, we investigated associations with intake of the corresponding food items in the UK Biobank using the same linear regression model.

Fourth, we quantified the variance explained for each food-liking trait in UK Biobank and ALSPAC, and for the corresponding food intake trait in UK Biobank, using the SNPs that were significantly associated with the food-liking trait in UK Biobank. We constructed a base model with only covariates (Formula 2) and a full model with covariates and SNPs associated with a trait (Formula 3). The variance explained for each trait was calculated as the difference between the R^2^ of the base model and that of the full model.

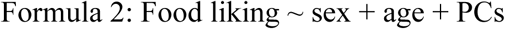

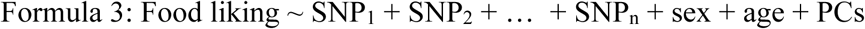

Where the SNPs were those significantly associated with a food-liking trait identified using formula 1.

Next, we selected SNPs to be used as IVs for dietary exposures. To ensure that a SNP only affects the outcome through one dietary exposure of interest, each food-liking associated SNP was screened based on: (i) it is only associated with one food-liking trait and its highly correlated traits (e.g., ice cream and cheesecake), and (ii) all food-liking associated SNPs in LD (r^2^ ≥ 0.1) with the selected SNP are also only associated with the same food-liking trait and its highly correlated traits. Given that MR is a method to assess the causal effect of long-term exposure, IVs were then tiered based on whether they have consistent effects across young (ALSPAC) and middle-aged adults (UK Biobank). To reflect the potential causal effect of food consumption, IVs were further tiered based on their association with food intake. Tier 1 IVs were SNPs with the same direction of effect on food liking in the ALSPAC (p-value < 0.05) and on food intake in the UK Biobank (p-value < 0.05); tier 2 SNPs were those with the same direction of effect on food liking in ALSPAC (p-value < 0.05); the remaining SNPs belonged to tier 3. See **Supplementary Figure 2** for a pipeline illustrating the selection of IVs and **Supplementary Table 1** for LD between SNPs.

Finally, we used SNPs significantly associated with food-liking traits as IVs in MR to assess the potential causal influences of food liking on cardiometabolic traits. SNP associations with cardiometabolic traits were extracted from published GWAS summary results statistics, including systolic blood pressure, diastolic blood pressure ^48^, high-density lipoprotein cholesterol, low-density lipoprotein cholesterol, total lipid cholesterol ^49^, fasting glucose, fasting insulin ^50^, BMI ^22^, coronary artery disease ^23^, and type 2 diabetes ^51^. All associations were extracted from GWAS primarily of European individuals. We prioritized using data without UK Biobank participants to avoid bias toward observational associations. See **Supplementary Table 2** for details and data sources of cardiometabolic traits.

MR analyses were performed using the Wald estimator when an exposure only had one IV, and the causal effect was calculated as *β̂_wald_* = *β̂_outcome_/β̂_exposure_* with the standard error being estimated using the Delta method as 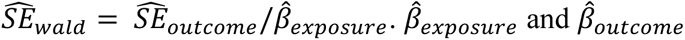 are the estimated effect of the IV on food liking and the cardiometabolic outcome, respectively; 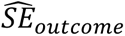 is the standard error of *β̂_outcome_*. When an exposure had two IVs, the causal effect was further calculated using the inverse variance-weighted method that meta-analyzes the *β̂_wald_* of each IV weighted by the inverse of their variance. Steiger filtering was applied to ensure that SNPs were consistent with the exposure-outcome direction; SNPs explaining more variance in the outcome than in the exposure would suggest reverse causation ^52^. For analyses with two IVs, we report Cochran’s Q as a heterogeneity check (df = 1). Pleiotropy-robust estimators that require three or more IVs (e.g., MR Egger ^53^, weighted median ^54^, MR-PRESSO ^55^) were not applicable. For significant MR results, if the outcome datasets contained UK Biobank participants, we performed sensitivity analyses to assess whether the effects were driven by UK Biobank by testing the IV-outcome association in UK Biobank using GWAS summary statistics results from the Neale lab (http://www.nealelab.is/uk-biobank/). See **Supplementary Note** for assumptions underlying Mendelian randomization and sensitivity analyses.

All analyses were performed using R version 4.4.1 and RStudio version 2024.4.1.748 (RStudio Team, 2020).

### Patient and public involvement

Patients and the public were not involved in the design, conduct, analysis, or reporting of this study. This research was based on secondary analysis of existing genetic and phenotypic data from population-based cohorts, and no new data were collected directly from participants. There are no plans to involve patients or the public in the dissemination of the results.

## Results

### Associations with food liking and intake

Of the 883 chemosensory receptor genes in the human genome (GRCh37/hg19), 30 taste receptor genes and 395 olfactory receptor genes are non-pseudo and have known chromosomal locations. Within these 425 genes, 7,400 single-nucleotide polymorphisms (SNPs) have a minor allele frequency ≥ 1%, of which 1,158 are missense, 29 are nonsense, and 27 are frameshift variants (**Supplementary Table 3**).

We tested the association between these 1,214 nonsynonymous SNPs and 140 food-liking traits in the UK Biobank. We identified 700 associations with an FDR-adjusted p-value < 0.05 across 268 SNPs within 117 genes (16 taste receptor genes and 101 olfactory receptor genes) and 96 food-liking traits (**Figure 2**; **Supplementary Figure 3**; **Supplementary Table 4**). The strongest associations were with liking garlic (p-value = 1.53 x 19^-69^ with *OR4K17* rs8005245), grapefruit (p-value = 1.88 x 10^-54^ with *TAS2R19* rs10772420), onions (p-value = 5.38 x 10^-41^ with *OR2T6* rs6587467), horseradish/wasabi (p-value = 2.28 x 10^-32^ with *TAS2R38* rs713598), adding salt to foods (p-value = 2.85 x 10^-24^ with *TAS2R38* rs10246939), and broad beans (p-value = 1.30 x 10^-19^ with *TAS2R16* rs860170). Most of these SNPs showed no association with the Townsend Deprivation Index in UK Biobank (p-value > 0.05; **Supplementary Table 4**), suggesting their effects on food liking were unlikely to be influenced by socioeconomic confounding factors. The few SNPs with nominal associations with the Townsend Deprivation Index were related to liking butternut squash, marzipan, plain yogurt, or strawberries.

**Figure 2:**
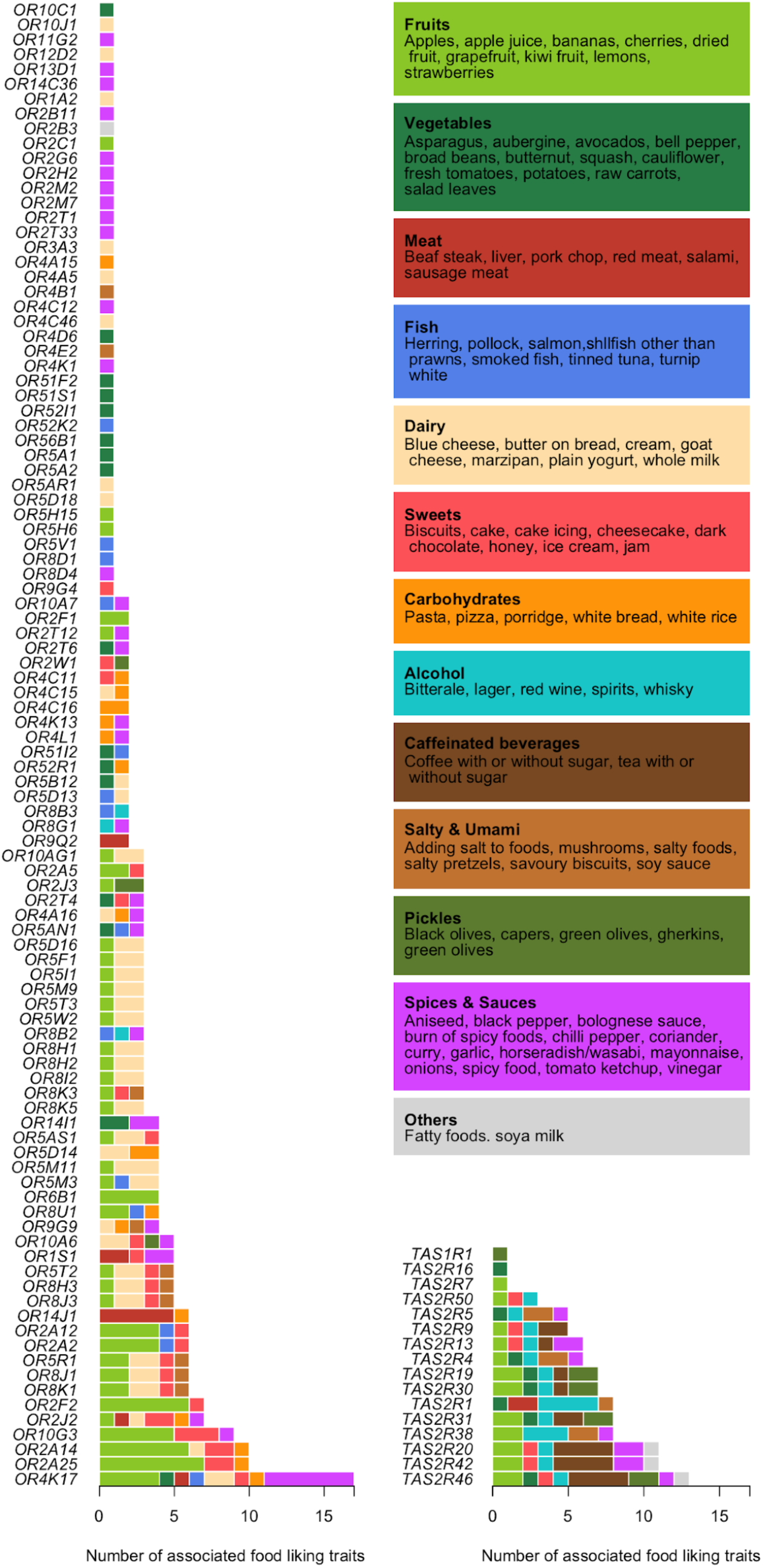
Associations between 96 food-liking traits and 268 nonsynonymous single-nucleotide polymorphisms (SNPs) within 101 olfactory (left panel) and 16 taste (right panel) receptor genes identified in UK Biobank (FDR-adjusted p-value < 0.05). Food items are clustered and colored according to their properties. A single SNP can be associated with up to 17 food-liking traits (i.e., *OR4K17* rs8005245), and 117 variants were only associated with one specific trait. Liking for fish, dairy, and carbohydrates is only related to variants within olfactory receptor genes, whereas liking for caffeinated and most alcoholic beverages is only associated with variants within taste receptor genes.

Among the 268 SNPs and 96 associated food-liking traits identified in UK Biobank, data for 223 SNPs or their proxy SNPs (i.e., SNPs in LD with r^2^ > 0.8; **Supplementary Table 5**) and 66 corresponding food-liking traits are available in ALSPAC (**Supplementary Table 6**). The majority of associations found in ALSPAC (410 out of 540 associations being tested) had the same direction of effect as observed in UK Biobank, among which 45 associations had a p-value < 0.05 (**Figure 3**, left panel); **Supplementary Table 4**). The strongest association identified in the UK Biobank (for liking garlic) was not significant in ALSPAC (p-value = 0.60 with *OR4K17* rs8005245).

**Figure 3:**
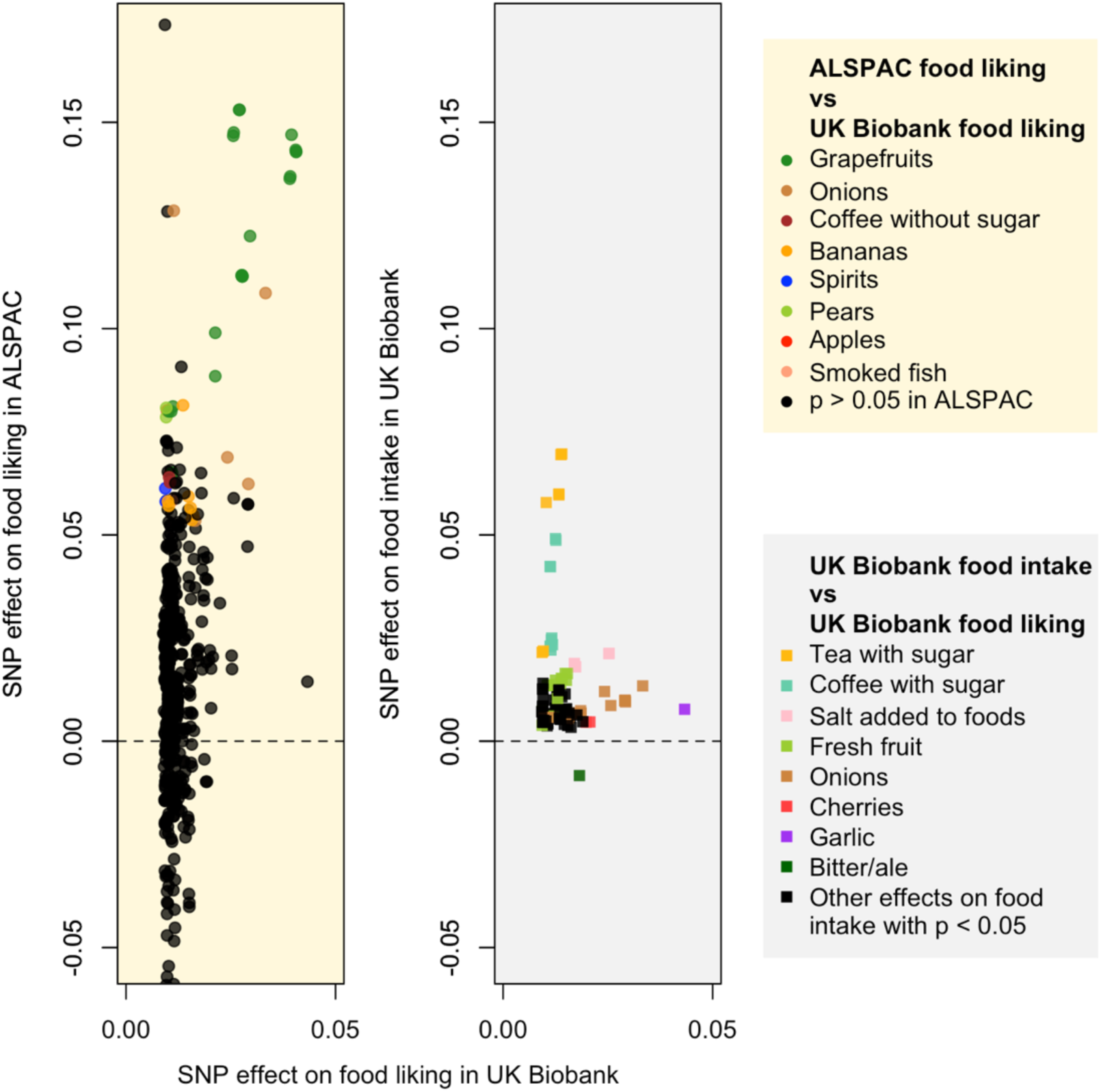
Comparison of standardized SNP effects on food-liking and intake traits in UK Biobank and Avon Longitudinal Study of Parents and Children (ALSPAC). **Left panel**: the most substantial concordant food-liking effects between UK Biobank and ALSPAC are for liking grapefruit and missense variants within a cluster of bitter taste receptors on chromosome 12 (*TAS2R31* rs10845293). Other concordant effects include variants for liking onions (*OR2T6* rs6587467), pears (*OR2A25* rs6951485), and apples (*OR2F2* rs2240359). There is a trend that the SNPs identified in the UK Biobank have a larger effect on the same food items in ALSPAC. **Right panel**: all food-liking SNPs affect their corresponding intake traits in the same direction; for example, the *OR4K17* rs8005245 C allele was associated with increased liking and higher intake of garlic (p-value = 7.17 x 10^-4^). The only exception is *TAS2R1* rs2234233 and bitter/ale, where the G allele is associated with increased liking and lower intake. This was probably because liking was measured for bitter or ale (both bitter), while intake was measured for beer or cider (with the latter being sweet). Only food-liking-associated SNPs associated with intake (p < 0.05) are plotted. Alleles are oriented to have a positive effect on food-liking traits.

We observed a trend that the variants identified in UK Biobank had a larger effect on the same food items in ALSPAC (**Figure 3**, left panel) and explained more variance (**Supplementary Figure 4**; **Supplementary Table 7**) in ALSPAC; however, cross-cohort heterogeneity tests showed no significant difference in effect estimates for most variants (p > 0.05; **Supplementary Table 4**). Among the 96 food-liking traits identified in UK Biobank, 60 had corresponding food-intake or proxied food-intake traits available in UK Biobank (**Supplementary Table 6**). Amongst the SNPs that were significantly associated with food liking, we identified 121 associations for intake of the same food items (p-value < 0.05), with 120 exhibiting the same direction of effect (**Figure 3**, right panel); **Supplementary Table 8**).

### Instrumental variable selection and Mendelian randomization

Among the 268 food-liking-associated SNPs, each was associated with 1 to 17 food-liking traits. After excluding SNPs (and SNPs in LD) with pleiotropic effects on multiple food-liking traits, we identified 28 IVs for 24 food-liking traits, including a tier 1 IV for liking onions (**Table 1**; see **Supplementary Table 9** for full results). The SNP *OR2T6* rs6587467 for liking onions had the same direction of effect on liking onions in UK Biobank (β = 0.087, p-value = 5.38 x 10^-41^, r^2^ = 0.11%) and ALSPAC (β = 0.23, p-value = 4.78 x 10^-4^, r^2^ = 0.48%), suggesting its long-lasting effect across adulthood. Further, the SNP was also associated with onion intake in the predicted direction (β = 0.002, p-value = 5.43 x 10^-9^, r^2^ = 0.02%).

**Table 1.** Instrumental variables for food-liking traits identified in the UK Biobank.

| Trait | Gene | SNP | EA/NEA | EAF | Beta | SE | P-value | r<sup>2</sup> | F-statistics | Tier* |
| --- | --- | --- | --- | --- | --- | --- | --- | --- | --- | --- |
| Onions | *OR2T6* | rs6587467 | G/T | 0.69 | 0.087 | 0.007 | 5.38 x 10<sup>-41</sup> | 0.11% | 179.627 | 1 |
| Bananas | *OR5H6* | rs9853906 | G/A | 0.69 | 0.029 | 0.007 | 3.91 x 10<sup>-5</sup> | 0.01% | 16.839 | 2 |
| Broad beans | *TAS2R16* | rs860170 | T/C | 0.68 | -0.08 | 0.009 | 1.30 x 10<sup>-19</sup> | 0.05% | 81.029 | 3 |
| Grapefruit | *TAS2R7* | rs619381 | T/C | 0.13 | 0.075 | 0.014 | 2.78 x 10<sup>-8</sup> | 0.02% | 30.740 | 3 |
| Tomato ketchup | *OR2B11* | rs12070953 | C/T | 0.14 | 0.057 | 0.012 | 2.74 x 10<sup>-6</sup> | 0.01% | 21.446 | 3 |
| Avocado | *OR10C1* | rs17177674 | A/C | 0.01 | -0.186 | 0.043 | 1.46 x 10<sup>-5</sup> | 0.01% | 18.344 | 3 |
| Vinegar | *OR4A16* | rs117538213 | C/T | 0.04 | 0.078 | 0.019 | 3.17 x 10<sup>-5</sup> | 0.01% | 17.161 | 3 |
| Cauliflower | *OR51I2* | rs12577167 | G/A | 0.12 | 0.038 | 0.009 | 3.90 x 10<sup>-5</sup> | 0.01% | 16.518 | 3 |
| Goats cheese | *OR1A2* | rs2469791 | T/C | 0.26 | -0.044 | 0.011 | 5.54 x 10<sup>-5</sup> | 0.01% | 16.236 | 3 |
| Salty pretzels | *OR4E2* | rs2874103 | A/G | 0.82 | -0.045 | 0.011 | 4.28 x 10<sup>-5</sup> | 0.01% | 16.226 | 3 |
| Garlic | *OR11G2* | rs4981088 | A/G | 0.52 | 0.028 | 0.007 | 1.59 x 10<sup>-4</sup> | 0.01% | 16.032 | 3 |
| Bananas | *OR2F2* | rs61739648 | A/G | 0.02 | 0.092 | 0.023 | 6.99 x 10<sup>-5</sup> | 0.01% | 15.867 | 3 |
| Black pepper | *OR8D4* | rs10790610 | C/T | 0.33 | 0.03 | 0.007 | 7.01 x 10<sup>-5</sup> | 0.01% | 15.800 | 3 |
| Ice cream | *OR2J2* | rs41270628 | T/C | 0.01 | -0.127 | 0.032 | 5.95 x 10<sup>-5</sup> | 0.01% | 15.750 | 3 |
| Green olives | *TAS1R1* | rs34160967 | A/G | 0.13 | -0.062 | 0.016 | 7.59 x 10<sup>-5</sup> | 0.01% | 15.595 | 3 |
| Fresh tomatoes | *OR56B1* | rs62621167 | T/C | 0.05 | 0.054 | 0.013 | 6.78 x 10<sup>-5</sup> | 0.01% | 15.578 | 3 |
| Aniseed | *OR13D1* | rs13294411 | G/C | 0.09 | -0.062 | 0.016 | 8.68 x 10<sup>-5</sup> | 0.01% | 15.439 | 3 |
| Goats cheese | *OR10J1* | rs12048482 | G/A | 0.64 | -0.039 | 0.01 | 8.92 x 10<sup>-5</sup> | 0.01% | 15.317 | 3 |
| Cream | *OR5M11* | rs628524 | T/C | 0.74 | 0.035 | 0.009 | 1.13 x 10<sup>-4</sup> | 0.01% | 14.893 | 3 |
| Apple juice | *OR2C1* | rs62000975 | A/G | 0.01 | 0.132 | 0.034 | 1.01 x 10<sup>-4</sup> | 0.01% | 14.87 | 3 |
| Plain yogurt | *OR3A3* | rs769432 | T/C | 0.59 | -0.032 | 0.008 | 1.12 x 10<sup>-4</sup> | 0.01% | 14.633 | 3 |
| Cheesecake | *OR2J2* | rs41270628 | T/C | 0.01 | -0.141 | 0.037 | 1.49 x 10<sup>-4</sup> | 0.01% | 14.228 | 3 |
| Pollock | *OR52K2* | rs61997232 | C/A | 0.03 | 0.098 | 0.026 | 1.52 x 10<sup>-4</sup> | 0.01% | 14.228 | 3 |
| Oranges | *OR5H15* | rs13082608 | T/C | 0.18 | -0.03 | 0.008 | 1.71 x 10<sup>-4</sup> | 0.01% | 14.085 | 3 |
| Shellfish other than prawns | *OR51I2* | rs35301588 | TCA/T | 0.38 | -0.041 | 0.011 | 1.62 x 10<sup>-4</sup> | 0.01% | 14.040 | 3 |
| Biscuits | *OR2T4* | rs57795102 | G/A | 0.01 | 0.129 | 0.035 | 1.86 x 10<sup>-4</sup> | 0.01% | 13.921 | 3 |
| Broad beans | *OR52I1* | rs146674516 | C/T | 0.02 | 0.116 | 0.031 | 2.03 x 10<sup>-4</sup> | 0.01% | 13.691 | 3 |
| Garlic | *OR4K1* | rs34394400 | T/C | 0.31 | -0.037 | 0.01 | 1.86 x 10<sup>-4</sup> | 0.01% | 13.626 | 3 |

\*Tier 1: The effect on food liking is replicated in ALSPAC (p-value < 0.05), and the SNP affects the intake of the same food in the UK Biobank (p-value < 0.05); Tier 2: The effect on food liking is replicated in the Avon Longitudinal Study of Parents and Children; Tier 3: others.
SNP, single nucleotide polymorphism; EA/NEA, effect allele/non-effect allele; EAF, effect allele frequency; SE, standard error.

MR showed that each point increased in the liking onions on a 9-point scale was associated with lower systolic blood pressure (β = -1.256 mmHg, p-value = 0.001), diastolic blood pressure (β = - 0.716 mmHg, p-value = 0.001), and the risk of type 2 diabetes (odds ratio [95% confidence interval] = 0.856 [0.781, 0.939], p-value = 0.001) (**Figure 4**). There was no strong evidence for a causal effect of liking onions on serum lipid cholesterol, blood glucose, BMI, or coronary artery disease. See MR results, Steiger filtering, and Cochrane’s Q tests for all food-liking traits in **Supplementary Table 10**.

**Figure 4:**
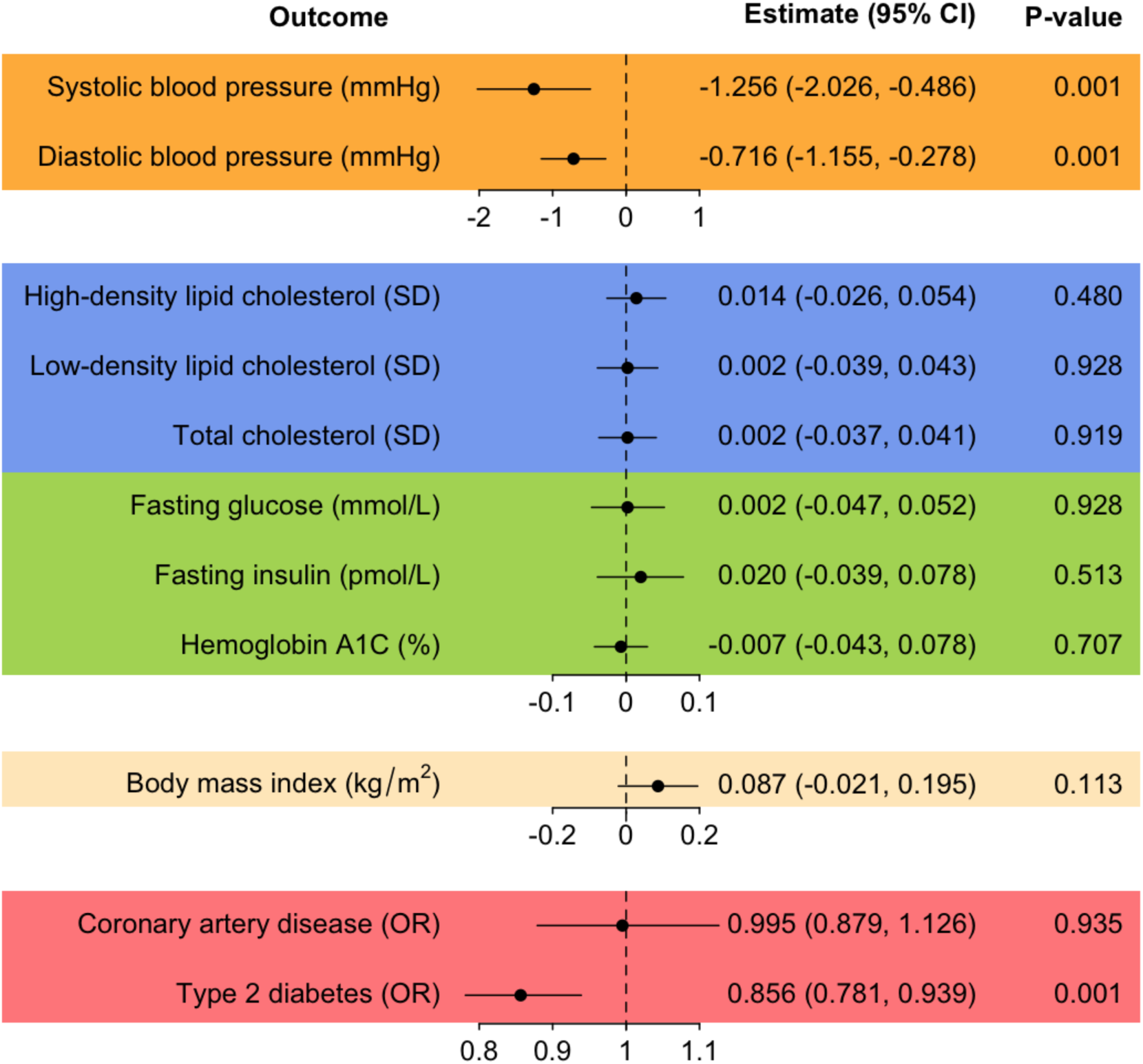
Causal effects of liking onions on cardiometabolic outcomes estimated using Mendelian randomization. Using *OR2T6* rs6587467 and the Wald estimator, we found that higher genetically predicted liking onions (i.e., one point increase on a 9-point scale) is associated with lower systolic blood pressure (β = -1.256 mmHg, p-value = 0.001), diastolic blood pressure (β = -0.716 mmHg, p-value = 0.001), and lower risk of type 2 diabetes (odds ratio [95% confidence interval] = 0.856 [0.781, 0.939], p-value = 0.001). There was no evidence of a causal relationship between onion intake and serum lipid cholesterol, blood glucose, body mass index, or coronary artery disease. SD, standard deviation; OR, odds ratio. The unit for hemoglobin A1C is expressed as the percentage (%) of total hemoglobin that is glycated.

## Discussion

This study sought to address a key methodological barrier in dietary MR: the lack of biologically credible IVs for dietary exposures. Many existing dietary instruments derive from significance-based GWAS conducted in older adults, in which diet may reflect underlying health rather than inherent preferences. We proposed that genetic variants for chemosensory perception—biological determinants of food liking established early in life—could serve as more robust instruments. Through a stringent multi-stage selection pipeline, we identified 28 instruments for 24 dietary exposures, including a high-confidence instrument for onion liking (*OR2T6* rs6587467) that demonstrated food-specificity, no association with socioeconomic status, and consistent effects across young- and late-adulthood cohorts. MR using this variant suggested a protective effect of onion consumption on lowering blood pressure and reducing the risk of type 2 diabetes, without detectable effects on BMI, glycaemic traits, or lipid levels.

Our approach, focusing on nonsynonymous variation within taste and olfactory receptor genes, identified SNPs within 16 taste and 101 olfactory receptor genes associated with 96 food-liking traits. These included 9 of the 10 non-pseudo chemosensory receptor genes previously identified in the GWAS of food liking using the same data set from UK Biobank ^19^. Many of these associations would not have been identified in a hypothesis-free GWAS using the conventional genome-wide significance threshold (i.e., p = 5 x 10^-8^). While some SNPs were linked to a single food, most variants were associated with multiple foods, consistent with the idea that individual receptors respond to shared volatiles, nutrients, or flavonoids across items ^56,57^. Most liking-associated SNPs also influenced intake of the corresponding foods in the same direction. Several associations were replicated in young adults from ALSPAC. This replication is important because the relationship between a genetic variant and food liking may change across the lifespan. The persistence of these associations from young adulthood into mid/late adulthood supports their validity as IVs for dietary exposures.

We intentionally restricted the screening to G protein-coupled receptors, i.e., the *TAS1R*/*TAS2R* families and *OR* repertoire, because these seven-transmembrane receptors directly mediate detection of tastants and odorants in the oral and nasal cavities ^58,59^. The *OR2T6* instrument is particularly compelling because *OR2T6* expression is restricted to the olfactory epithelium ^60,61^, providing biological coherence for its role in onion perception, preference, and intake. If the observed association between onion intake and cardiometabolic outcomes reflects a causal relationship, it could be explained by onions’ bioactive properties, including flavonoids, organosulfur compounds, and antioxidants ^62^. For instance, small clinical studies have reported that eating red onions lowers blood sugar in diabetic patients ^63^, and quercetin, a flavonoid found in onions, has been shown to modulate pancreatic islet function and insulin release in rodent models ^64^. A possible hypothesis is that onion consumption may have greater metabolic benefits in individuals with prediabetes or early insulin resistance than in metabolically healthy individuals. These findings warrant replication in independent cohorts and mechanistic investigation before informing dietary recommendations.

The present study has limitations. The *TAS1R*, *TAS2R*, and *OR* gene families do not include genes underlying the salty or sour taste modalities, which are mediated predominantly by ion channels rather than G protein-coupled receptors ^65^. Some chemosensory receptors are also expressed outside the oral-nasal cavity and may have additional physiological functions ^66^, potentially introducing pleiotropic effects. Selection bias is another potential issue, as the UK Biobank has a response rate of ∼5% and participants differ from the underlying population in socioeconomic status, health, and survival ^67^. The 24-hour recall and food-liking subsamples are more restricted, and the same applies to the ALSPAC food-liking subsample. If study participation depends jointly on genotype and phenotype, MR estimates may be biased. Therefore, replication in larger, more diverse, and more representative cohorts will be necessary for validation.

Despite these limitations, our findings demonstrate the potential of chemosensory genetics to strengthen causal inference in nutrition research. Chemosensory variants offer biologically grounded IVs that can reduce pleiotropy and reverse causation, thereby addressing long-standing limitations in dietary MR. The instrument for onion liking illustrates the feasibility and utility of this approach. Future work should extend this framework to additional chemosensory pathways and develop analytical methods that jointly model multiple, correlated foods to improve causal estimation of dietary patterns.

## Data Availability

All data produced in the present study are available upon reasonable request to the authors.

## Acknowledgement

We are extremely grateful to all the families who participated in this study, the midwives for their help recruiting them, and the ALSPAC team, including interviewers, computer and laboratory technicians, clerical workers, research scientists, volunteers, managers, receptionists, and nurses. We thank those who contributed to the survey design and data collection of food preferences and intake from UK Biobank.

LDH, DRR, and PVJ designed research; LDH and PVJ accessed data; LDH and CL analyzed data and created figures; LDH wrote the initial manuscript; all authors contributed to data analyses and critically reviewed the manuscript. LDH had primary responsibility for final content. All authors read and approved the final manuscript.

## Data Availability

The UK Biobank data (https://www.ukbiobank.ac.uk/) were accessed with application ID 57780. The ALSPAC data (https://www.bristol.ac.uk/alspac/) were accessed with application ID B3544. GWAS summary statistics results of the Townsend Deprivation Index are available in the OpenGWAS database (https://gwas.mrcieu.ac.uk/). GWAS summary statistics results from the Neale lab were accessed at (http://www.nealelab.is/uk-biobank/). The analysis pipeline code is available at https://github.com/Cailu086Lin/UKBioBank_Data_Processing_TR_OR_Food_Liking and https://github.com/danielldhwang/ALSPAC_Food_Liking

