## Supplementary Materials for "Biologically informed instrument selection for dietary Mendelian randomization using chemosensory receptor variants"

**Supplementary Figure 1**. Overview of the analyses performed in the present study. ALSPAC, Avon Longitudinal Study of Parents and Children.

**Supplementary Figure 2.** Pipeline showing instrumental variables selection and tiering for Mendelian randomization.

**Supplementary Figure 3.** 700 significant associations between food-liking traits and nonsynonymous single nucleotide polymorphisms (SNPs) within chemosensory receptor genes identified in UK Biobank (FDR-adjusted p-value < 0.05).

**Supplementary Figure 4.** Variance explained in food-liking and intake traits by SNPs within chemosensory receptor genes.

**Supplementary Note**

*Mendelian randomization (MR)*

*Sensitivity analyses of the effects of onion intake on blood pressure and type 2 diabetes*

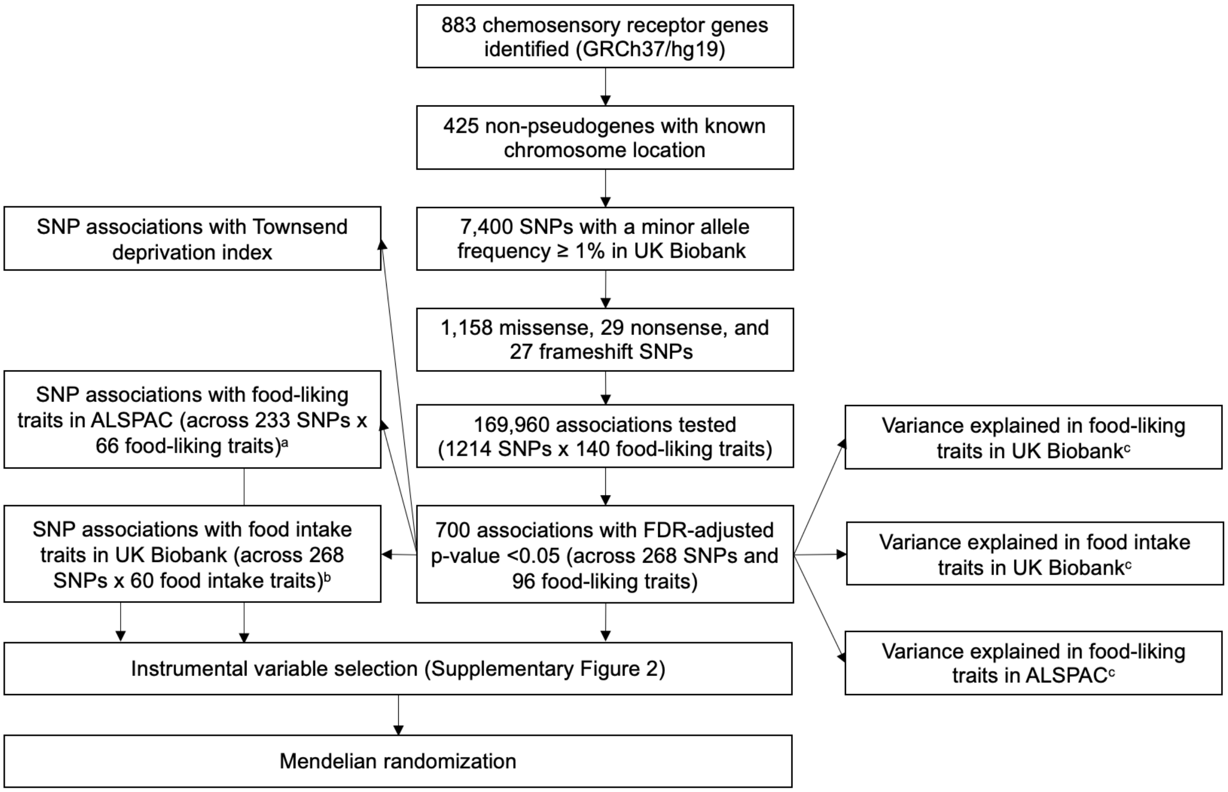


**Supplementary Figure 1**. **Overview of the analyses performed in the present study.** ALSPAC, Avon Longitudinal Study of Parents and Children. SNP, single nucleotide polymorphism. [a]Among 268 SNPs and 99 food-liking traits identified in the UK Biobank, 233 SNPs and 66 food-liking traits were available in the ALSPAC. [b]Among 96 food-liking traits in UK Biobank, 60 had their corresponding food intake traits available in UK Biobank. [c]Variance explained in each trait by SNPs associated with their corresponding food-liking traits identified in UK Biobank.

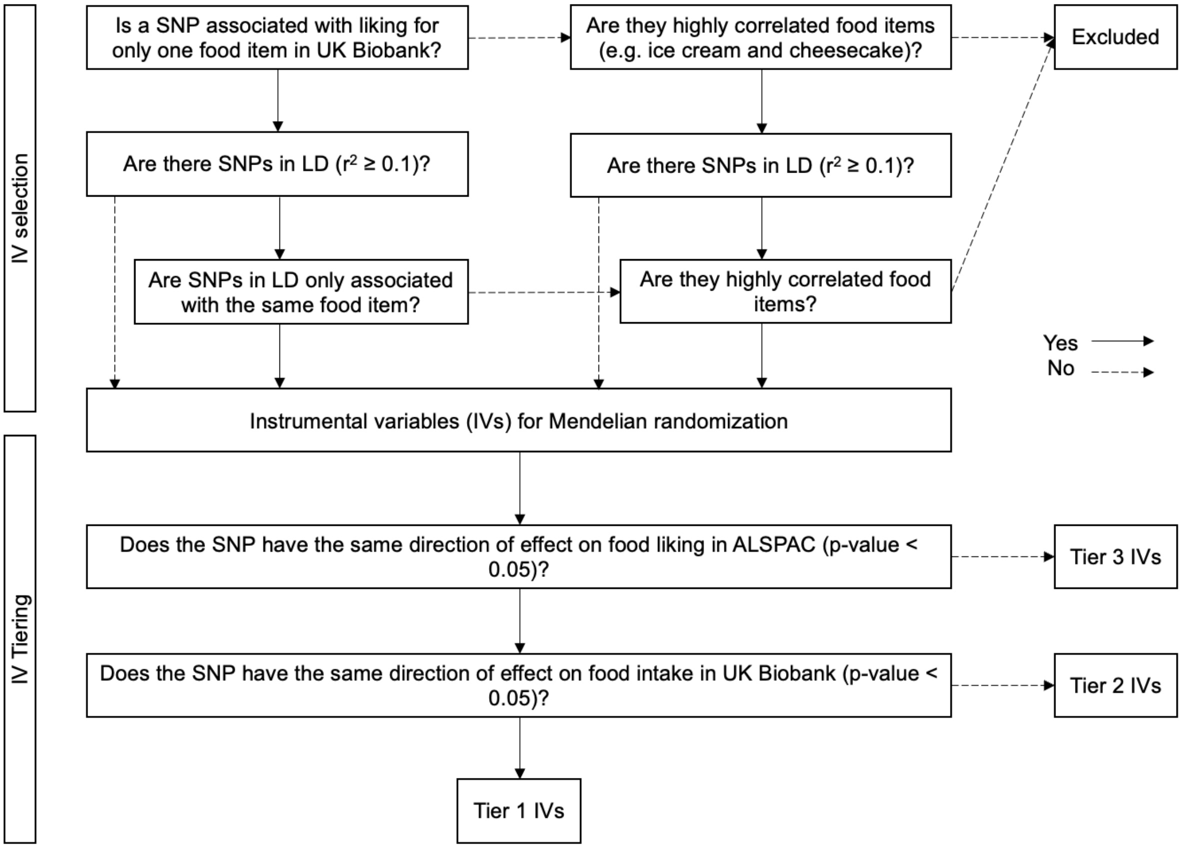


**Supplementary Figure 2. Pipeline showing instrumental variables selection and tiering for Mendelian randomization.** SNP, single nucleotide polymorphism; LD, linkage disequilibrium; IVs, instrumental variable; ALSPAC, Avon Longitudinal Study of Parents and Children.

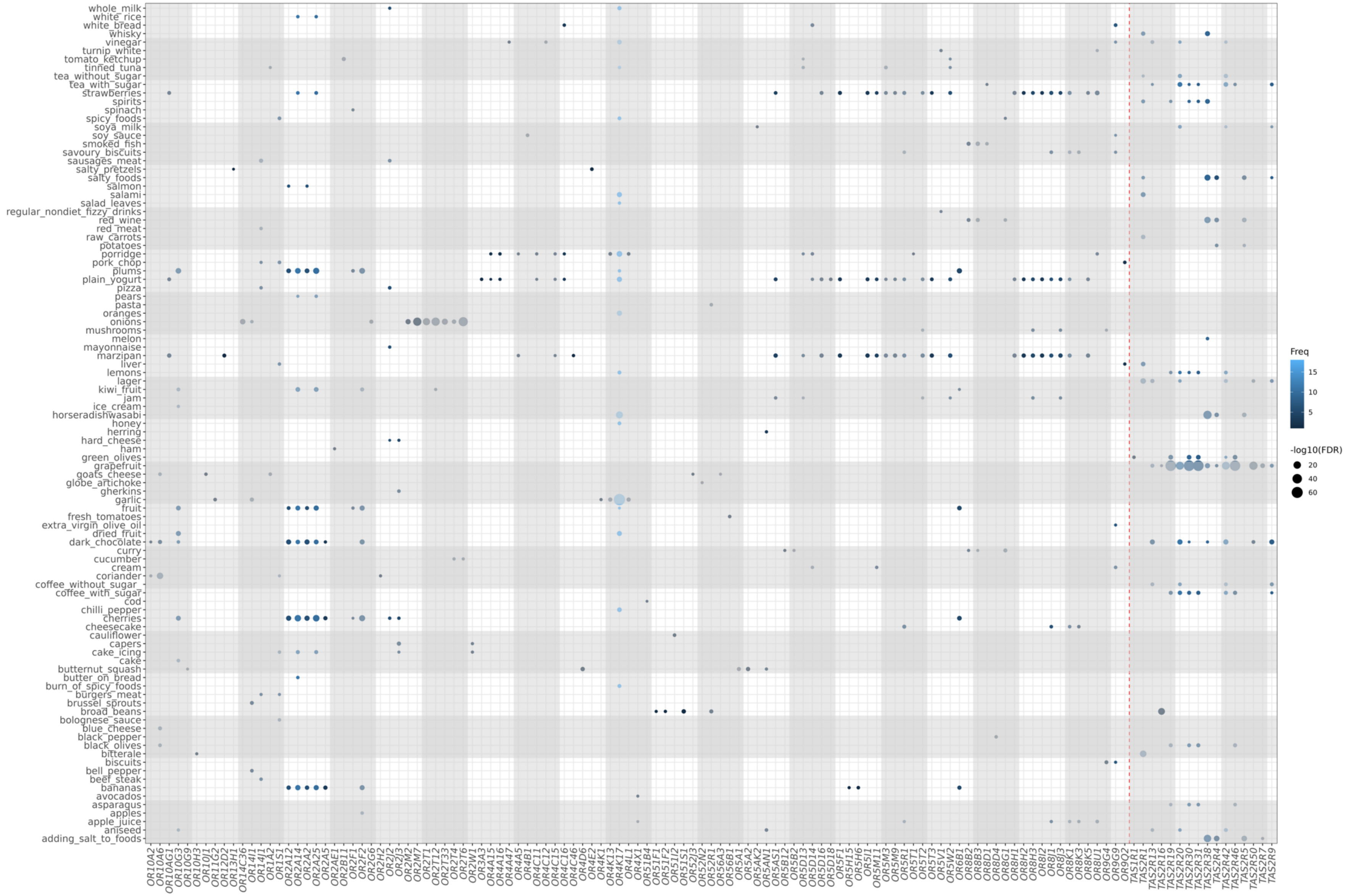


**Supplementary Figure 3. 700 significant associations between food-liking traits and nonsynonymous single nucleotide polymorphisms (SNPs) within chemosensory receptor genes identified in UK Biobank (FDR-adjusted p-value < 0.05).** The frequency (Freq) is the sum of associations between all SNPs within a gene and their associations with food-liking traits.

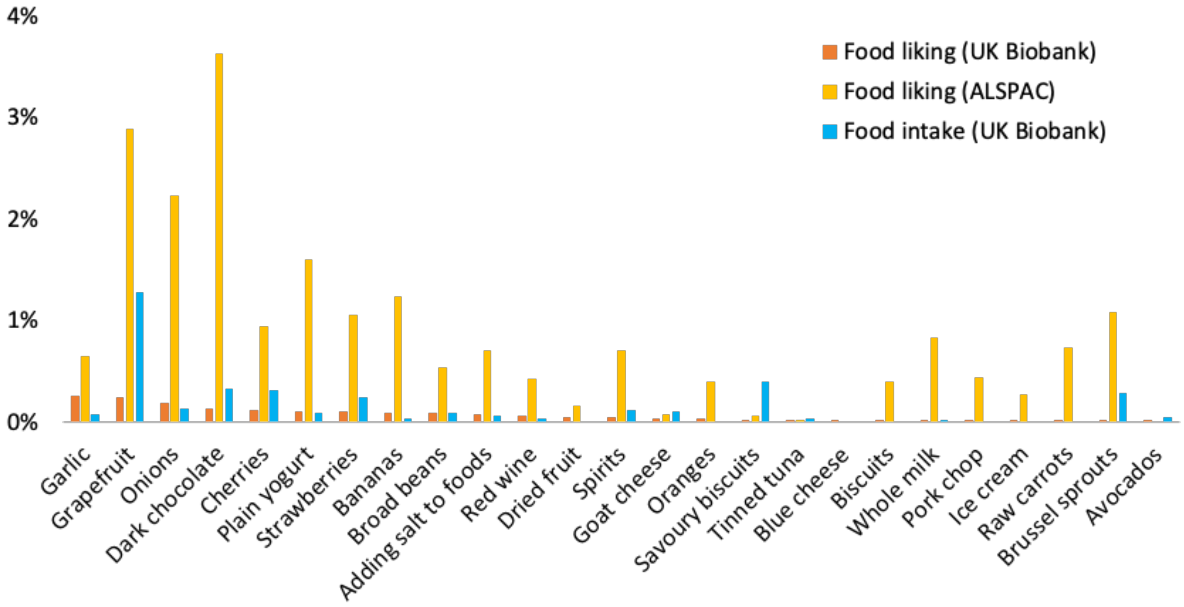


**Supplementary Figure 4. Variance explained in food-liking and intake traits by SNPs within chemosensory receptor genes.** For the 96 food-liking traits in UK Biobank with significantly associated SNPs, their correspondingly associated variants explained > 0.1% of the variance in 10 food-liking traits, which were liking garlic (0.26%), grapefruit (0.24%), onions (0.19%), horseradish/wasabi (0.16%), dark chocolate (0.13%), cherries (0.11%), plain yogurt (0.11%), plums (0.11%), coriander (0.11%), and strawberries (0.11%). In ALSPAC, the same set of variants explained > 0.1% of the variance in 47 food-liking traits, with the highest being dark chocolate (3.63%), followed by grapefruit (2.89%), onions (2.23%), plain yogurt (1.60%), chili pepper (1.41%), bananas (1.23%), marzipan (1.22%), Brussel sprouts (1.09%), and strawberries (1.06%). For the corresponding food intake traits in UK Biobank, the same set of variants explained > 0.1% of the variance in 18 of them, which were grapefruit (1.28%), artificial sweetener added to tea (0.61%), artificial sweetener added to coffee (0.43%), savory biscuits (0.40%), dark chocolate (0.32%), cherry (0.32%), sprouts (0.28%), berry (0.25%), lobster/crab (0.23%), olives (0.20%), porridge (0.16%), plum (0.14%), onion (0.14%), spirits (0.13%), salted nuts (0.12%), other tea (0.12%), other fish (0.11%), and cheesecake (0.11%). Only the top 25 food traits in both UK Biobank and ALSPAC are presented here.

**Supplementary Note**

*Mendelian randomization (MR)*

MR is an epidemiological method that uses genetic variants robustly associated with a modifiable exposure as instrumental variables (IVs) to assess a potential causal relationship between an exposure and an outcome of interest and to estimate the magnitude of this causal effect. MR can be considered analogous to a randomized controlled trial, but it involves individuals' genotypes rather than treatments. In MR, the segregation of alleles during meiosis is similar to the randomization process in RCTs, except that in MR, the randomization occurs at meiosis, and the causal effects estimated in the analysis represent the long-term effects of life-long exposures. Mendel's Law of Segregation ensures that genetic variants segregate randomly and independently of environmental factors. At the same time, Mendel's Law of Independent Assortment suggests that genetic variants should also segregate independently of other traits in most cases. This means genetic variants are less susceptible to confounding than the variables used in "traditional" observational epidemiological studies.

The three MR core assumptions for SNPs to be used as valid IVs are: i) SNPs are robustly associated with the exposure (the relevance assumption), ii) SNPs are not associated with factors that confound the association between the exposure and the outcome (the independence assumption), and iii) SNPs influence the outcome solely through their effect on the exposure (the exclusion restriction assumption), as illustrated in the below figure.

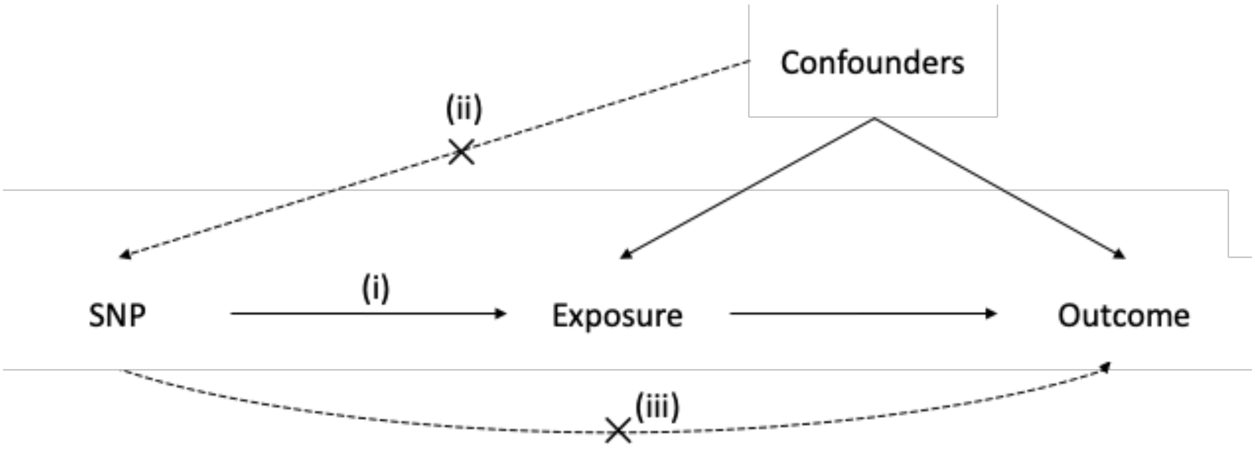


**Three core assumptions of Mendelian randomization**

To address the first MR assumption, we calculated F-statistic for each SNP. All SNPs had F-statistics > 10 and were considered strong IVs. To address the second assumption, which may be violated by population stratification, we limited the data used in MR to GWAS which included individuals of only/predominantly European ancestry and genetic principal components as covariates. The third assumption is addressed using only SNPs with chemosensory receptor genes to instrument food-liking exposures.

*Sensitivity analyses of the effects of liking onions on blood pressure and type 2 diabetes*
Given that the significant effects of liking onions were identified using outcome datasets containing UK Biobank participants (i.e., GWAS meta-analysis including UK Biobank), we performed sensitivity analyses using outcome data only from UK Biobank to investigate whether the effects were driven by sample overlap between the exposure and the outcome. The IV-outcome associations in the UK Biobank European participants were extracted from the GWAS summary results statistics from the Neale lab (http://www.nealelab.is/uk-biobank/).

| | Outcome (unit) | Data source | Sample size | SNP effect on outcome | | MR estimate of 1 unit increase in onion liking (95% CI) |
|---|---|---|---|---|---|---|
| | | | | Beta/OR (95% CI) | P-value | |
| Primary analysis | SBP (mmHg) | Evangelou (2018) | 757,601 | -0.109 (-0.176, -0.041) | $1.38 \times 10^{-3}$ | -1.256 (-2.026, -0.486) |
| | DBP (mmHg) | Evangelou (2018) | 757,601 | -0.062 (-0.101, -0.024) | $1.39 \times 10^{-3}$ | -0.716 (-1.155, -0.278) |
| | T2D (Odds ratio) | Suzuki (2024) | 1,812,017 (13% cases) | 0.987 (0.979, 0.994) | $1.07 \times 10^{-3}$ | 0.856 (0.781, 0.939) |
| Sensitivity analysis | SBP (mmHg) | Neale lab (UK Biobank) | 340,159 | -0.087 (-0.177, 0.002) | 0.056 | -1.000 (-2.034, 0.023) |
| | DBP (mmHg) | Neale lab (UK Biobank) | 340,168 | -0.035 (-0.087, 0.016) | 0.175 | -0.402 (-1, 0.184) |
| | T2D (Odds ratio) | Neale lab (UK Biobank) | 361,194 (4% cases) | 0.999 (0.0999, 1.000) | 0.736 | 0.999 (0.997, 1.002) |

The results showed that the IV for liking onions (*OR2T6* rs6587467) was not associated with systolic blood pressure (SBP), diastolic blood pressure (DBP), or type 2 diabetes (T2D) risk in UK Biobank (p-value > 0.05), and the MR results were null. Furthermore, the point estimates of the SNP effects on the outcomes in UK Biobank were smaller than in the GWAS meta-analyses. These results support that the significant effects of liking onions were not driven by sample overlap.
